# Metabolic Polygenic Risk Scores for Prediction of Obesity, Type 2 Diabetes, and Related Morbidities

**DOI:** 10.1101/2024.11.01.24316620

**Authors:** Min Seo Kim, Qiuli Chen, Yang Sui, Xiong Yang, Shaoqi Wang, Lu-Chen Weng, So Mi Jemma Cho, Satoshi Koyama, Xinyu Zhu, Kang Yu, Xingyu Chen, Rufan Zhang, Wanqing Yin, Shuangqiao Liao, Zhaoqi Liu, Fowzan S Alkuraya, Pradeep Natarajan, Patrick T. Ellinor, Akl C. Fahed, Minxian Wang

## Abstract

Obesity and type 2 diabetes (T2D) are metabolic diseases with shared pathophysiology. Traditional polygenic risk scores (PRS) have focused on these conditions individually, yet the single disease approach falls short in capturing the full dimension of metabolic dysfunction. We derived biologically enriched metabolic PRS (MetPRS), a composite score that uses multi-ancestry genome-wide association studies of 22 metabolic traits from over 10 million people. MetPRS, optimized to predict obesity (O-MetPRS) and T2D (D-MetPRS), was validated in the UK Biobank (UKB, n=15,000), and tested in UKB hold-out set (n=49,377), then externally tested in 3 cohorts – All of Us (n=245,394), Mass General Brigham (MGB) Biobank (n=53,306), and a King Faisal Specialist Hospital and Research Center cohort (n=6,416). O-MetPRS and D-MetPRS outperformed existing PRSs in predicting obesity and T2D across 6 ancestries (European, African, East Asian, South Asian, Latino/admixed American, and Middle Eastern). O-MetPRS and D-MetPRS also predicted morbidities and downstream complications of obesity and T2D, as well as the use of GLP-1 receptor agonists in contemporary practice. Among 37,329 MGB participants free of T2D and obesity at baseline, those in the top decile of O-MetPRS had a 103% relatively higher chance, and those in the top decile of D-MetPRS had an 80% relatively higher chance of receiving a GLP-1 receptor agonist prescription compared to individuals at the population median of MetPRS. The biologically enriched MetPRS is poised to have an impact across all layers of clinical utility, from predicting morbidities to informing management decisions.

## Introduction

Obesity and type 2 diabetes (T2D) are closely interconnected through a shared pathophysiology, often characterized by insulin resistance, and together constitute a major portion of metabolic diseases. This relationship, frequently referred to as "diabesity",^1^ reflects the overlapping genetic architecture of both conditions, emphasizing the need to study their combined effects on metabolic health and related morbidities.^2,3^ While research into the shared and distinct genetic bases of metabolic traits has provided valuable biological insights,^4^ this multivariable knowledge has not been fully translated into clinical utility via genetic risk assessment. The polygenic risk score (PRS) has emerged as a powerful tool for assessing composite genetic risks, gaining traction for its potential clinical applications.^5,6^

The PRSs for obesity and T2D have been developed individually based on a single disease approach thus far,^7^ without accounting for the multivariable nature of these conditions. We and others previously demonstrated that incorporating genetic information for relevant diseases enabled a significant boost in disease risk predictive performance.^8–10^ Building on this multi-disease approach, we have now further incorporated a broader spectrum of measures for diabesity, acknowledging that different measures of obesity (e.g., body mass index[BMI], waist circumference[WC], waist-to-hip ratio[WHR]) and diabetes (e.g., HbA1c, fasting glucose[FG], insulin sensitivity) capture intersecting but also complementary biology of these conditions. For instance, BMI typically measures overall body mass, whereas WC provides more specific information about abdominal or central adiposity.^11^ Leveraging multiple indices that measure various dimensions of metabolic dysfunction may effectively capture the underlying metabolic complexity and potentially translate it into a biologically enriched and clinically meaningful PRS.

By integrating genetic information for related diseases and measures, we developed a metabolic PRS (MetPRS) optimized for obesity (O-MetPRS) and T2D (D-MetPRS). We incorporated genetic data from 22 metabolic traits, utilizing multi-ancestry genome-wide association study (GWAS) data from over 10 million participants. The model was validated and tested in the UK Biobank (UKB), and externally tested in three multiethnic cohorts comprising up to 300,000 participants. This multi-disease and multi-measure approach has resulted in a biologically enriched and cross-ancestry transferable PRS capable of predicting a broad spectrum of metabolic health outcomes with high accuracy and relevance to clinical care.

## Results

We integrated GWAS summary statistics from 22 obesity and diabetes related traits including, but not limited to, BMI, body weight, body fat percentage, WC, WHR, hip circumference, abdominal subcutaneous adipose tissue (ASAT), visceral adipose tissue (VAT), gluteofemoral adipose tissue (GFAT), HbA1c, fasting glucose, fasting insulin, 2-hr glucose, and insulin sensitivity index (**Supplementary Table 1**). The GWAS summary statistics were derived from the largest contemporary data consortia, including Million Veteran Program (MVP), FinnGen, Biobank Japan (BBJ), UK Biobank (UKB), and Taiwan Biobank, encompassing over 10 million participants. Participants in the UK Biobank were divided into three distinct groups: the dataset for GWAS, the training set, and the hold-out test set, with no overlap between samples across these groups (**Extended Data Fig.1**). The obesity and T2D models were validated on 15,000 individuals of European ancestry, including 3,814 individuals with obesity (BMI ≥ 30 Kg/m^2^ for European) and 1,196 with T2D. UKB hold-out test set consisted of 49,377 individuals, with 70.1% of European ancestry, 13.9% of African ancestry, 2.6% of East Asian ancestry, and 13.4% of South Asian ancestry (**Extended Data Fig.1 and Supplementary Table 2**). The remaining European-ancestry samples (N=378,921) were used for *de novo* GWAS analysis, and incorporated in the PRS score development (**Supplementary Table 1**). The model development process is illustrated in **Fig. 1**.

**Fig 1.**
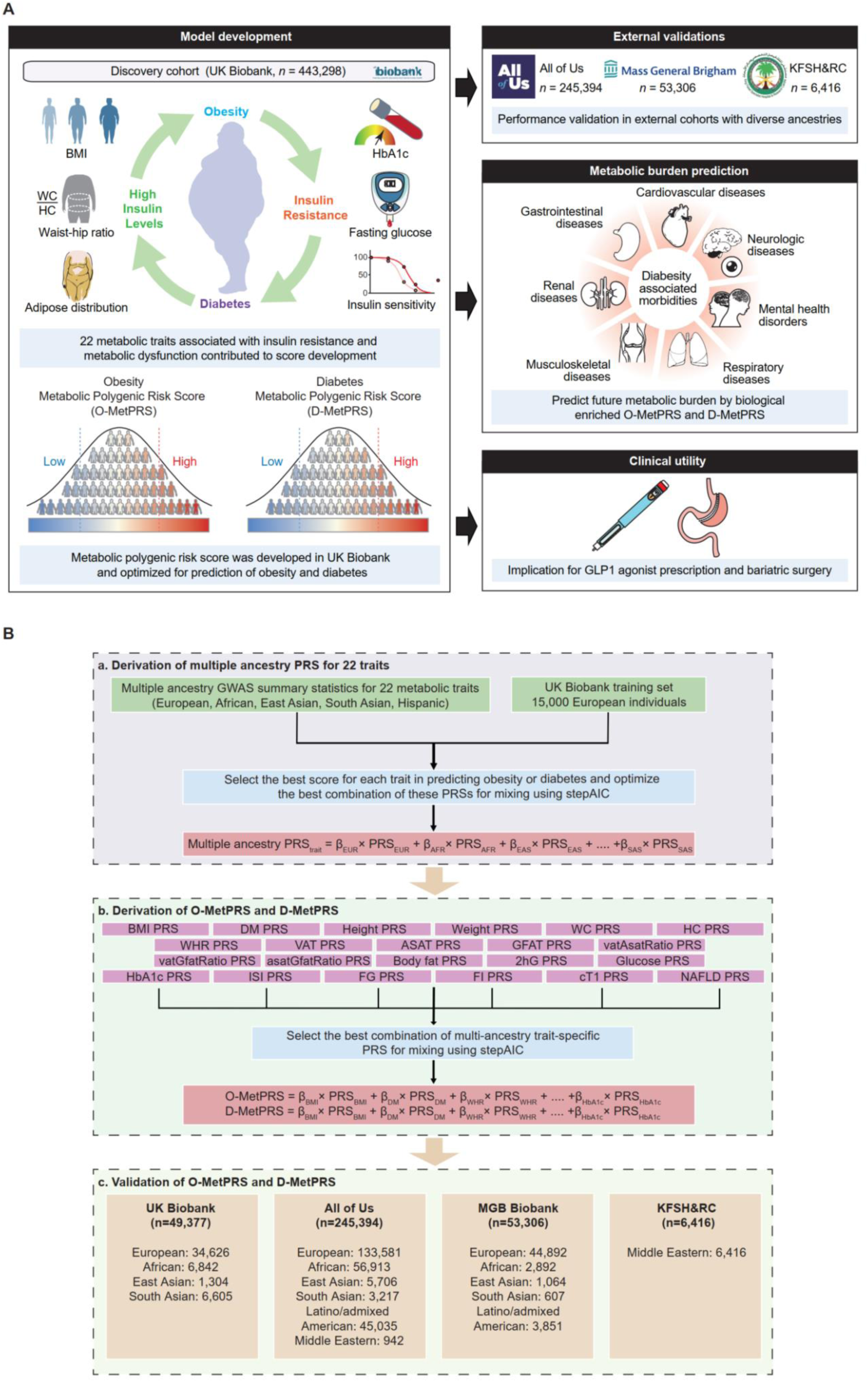
Study overview. Overview of study **(A)** and MetPRS development **(B)**. Polygenic risk scores were developed using cohort-specific, ancestry-stratified summary statistics for 22 metabolic traits. (a) For each trait, such as BMI, the optimal combination of cohort-specific, ancestry-stratified PRSs was identified using stepAIC, the selected PRSs were then combined linearly to create multi-ancestry scores for predicting target trait (Obesity or Type 2 diabetes). The best mixing weights (β) were calculated using regression on 15,000 European participants from the UK Biobank training set (layer 1).(b) The most effective combination of multi-ancestry, trait-specific PRSs was identified using stepAIC for predicting target trait, and their optimal mixing weights (β) for selected PRSs were determined through regression on the same dataset. The chosen PRSs were then linearly combined to produce O-MetPRS and D-MetPRS (layer 2). The participants in the validation and testing set are non-overlapping individuals and independent with input GWAS samples. (c) These MetPRS were tested on internal (UK Biobank hold out test set) and external (All of Us, MGB Biobank, KFSH&RC) datasets. The genetic predispositions to traits indicative of obesity (i.e., BMI, waist circumference, and visceral adipose tissue) or T2D (i.e., fasting glucose, HbA1c, and insulin sensitivity) can contribute unique insights into the multifaceted nature of metabolic dysfunction. Leveraging genetic signature of related traits might capture a more comprehensive underlying metabolism and enhance the overall prediction of obesity and T2D. BMI: body mass index; DM: type 2 diabetes; WC: waist circumference; HC: hip circumference; WHR: waist-to-hip ratio; VAT: visceral adipose tissue; ASAT: abdominal subcutaneous adipose tissue; GFAT: gluteofemoral adipose tissue; vatGfatRatio: VAT/GFAT ratio; asatGfatRatio: ASAT/GFAT ratio; vatAsatRatio: VAT/ASAT ratio; 2hG: 2-hour glucose; ISI: insulin sensitivity index; FG: fasting glucose; FI: fasting insulin; cT1: MRI-based Liver iron corrected T1; NAFLD: non-alcoholic fatty liver disease.

### Genetic Correlations Among Metabolic Traits

We selected 22 traits associated with diabesity and insulin resistance for which GWAS summary statistics are currently available and have no sample overlap with UK biobank participants (**Supplementary Table 1)**. We assessed the genetic correlations among 22 metabolic traits using cross-trait LD-score regression (**Fig. 2**). Significant correlations were observed between traits associated with obesity, such as BMI, WC, body weight, VAT, reflecting their shared genetic underpinnings. For example, the genetic correlation between BMI and WC was 0.90, while VAT showed strong correlations with both body fat (0.71) and WC (0.79), which remained significant after multiple testing corrections (**Fig. 2** and **Supplementary Table 3**). In contrast, the Modified Stumvoll insulin sensitivity index (ISI)^12^ displayed negative genetic correlations with most traits, as the index is inversely related to insulin resistance.

**Fig. 2.**
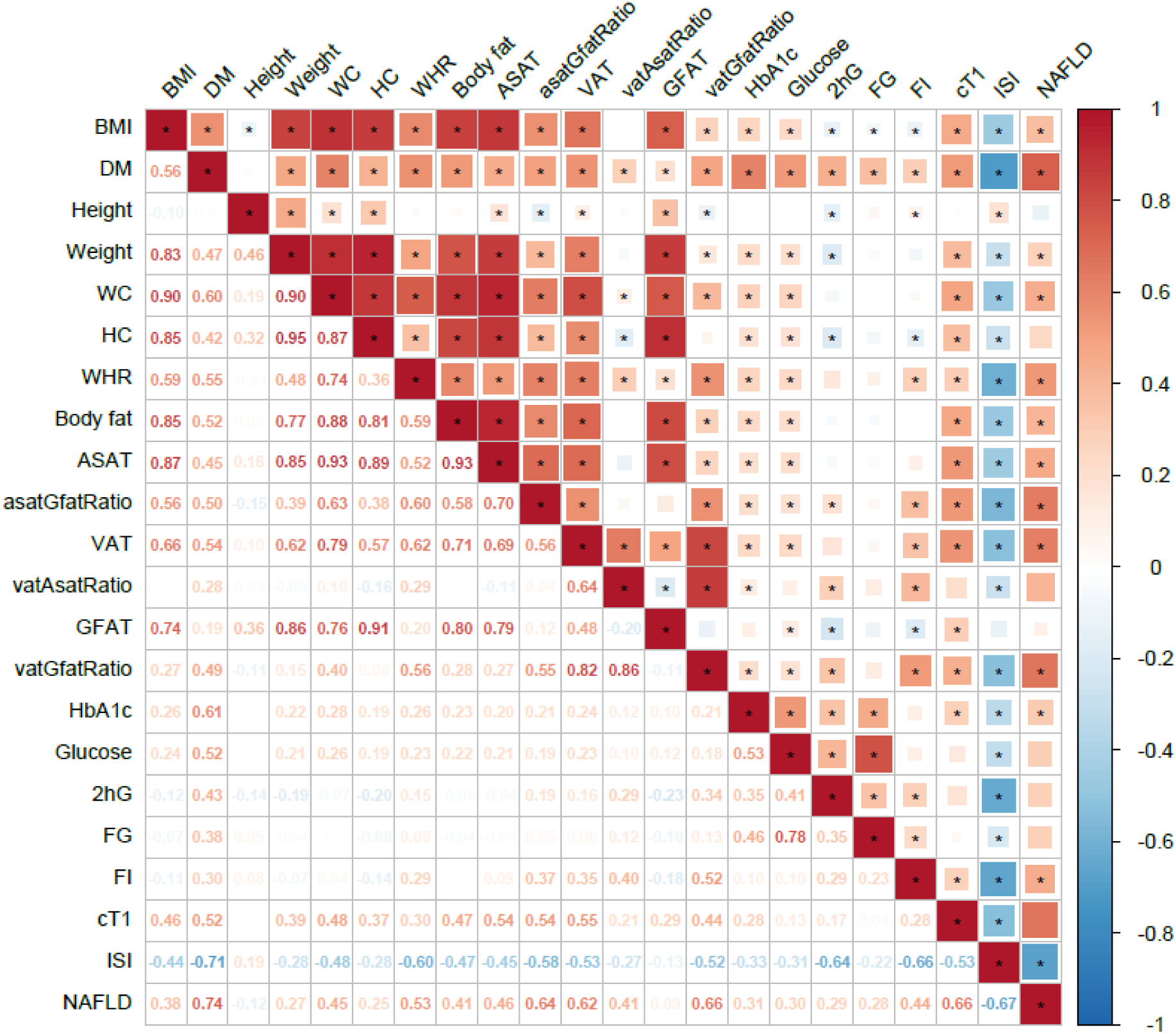
Genetic correlations across 22 metabolic traits and measures associated with insulin resistance. Genetic correlations were obtained from cross-trait LD-score regression using sex-combined summary statistics. Stumvoll insulin sensitivity index (ISI) is inversely proportional to insulin resistance as it measures insulin sensitivity. Genetic correlation heatmap matrix remained significant after multiple testing corrections are indicated by asterisks. BMI: body mass index; DM: type 2 diabetes; WC: waist circumference; HC: hip circumference; WHR: waist-to-hip ratio; ASAT: abdominal subcutaneous adipose tissue; VAT: visceral adipose tissue; GFAT: gluteofemoral adipose tissue; asatGfatRatio: ASAT/GFAT ratio; vatAsatRatio: VAT/ASAT ratio; vatGfatRatio: VAT/GFAT ratio; 2hG: 2-hour glucose; FG: fasting glucose; FI: fasting insulin; cT1: MRI-based Liver iron corrected T1; ISI: insulin sensitivity index; NAFLD: non-alcoholic fatty liver disease.

### Model Development and Internal Validation of MetPRS in the UK Biobank

We used a previously described multi-ancestry and multi-trait GWAS integration framework^8^ for developing high-performance and cross-ancestry transferable metabolic PRS (MetPRS), with the purpose specifically optimized to predict obesity (O-MetPRS) and T2D (D-MetPRS), **Fig 1**. The predictive performance of O-MetPRS and D-MetPRS was first evaluated in the hold-out test set from the UK Biobank (n = 49,377) (**Fig. 3 and Extended Data Fig.1**). For obesity among European participants, O-MetPRS achieved an odds ratio per standard deviation (OR/SD) of 2.22 (95% CI: 2.16–2.28), outperforming all previously published PRSs for obesity – the second best performing PRS for obesity (PGS catalog: PGS000027) had an OR/SD of 1.79 (95% CI: 1.75–1.84) (**Fig. 3A and Supplementary Table 4**). Moreover, the range of obesity prevalence stratified by O-MetPRS was significantly broader, spanning from 2.9% to 70.3% across percentiles, compared to 6.3% to 58.2% for the second best performing PRS, PGS000027 (**Fig. 3B**). For T2D, D-MetPRS showed an OR/SD of 2.08 (95% CI: 2.00–2.18), also exceeding the performance of all previously published PRSs (**Fig. 3C and Supplementary Table 5**). Similarly, D-MetPRS stratified a notably wider range of T2D prevalence, from 0.9% to 28.8% across percentiles, compared to 2.6% to 23.9% for the second best PGS001357 (**Fig. 3D**). Likewise, O-MetPRS and D-MetPRS consistently ranked as the top-performing PRSs for obesity and T2D among African and East Asian participants (**Fig. 3A and Fig. 3C**). These findings demonstrate that MetPRS provides superior risk stratification for both obesity and T2D in diverse ethnic groups compared to existing PRSs.

**Fig 3.**
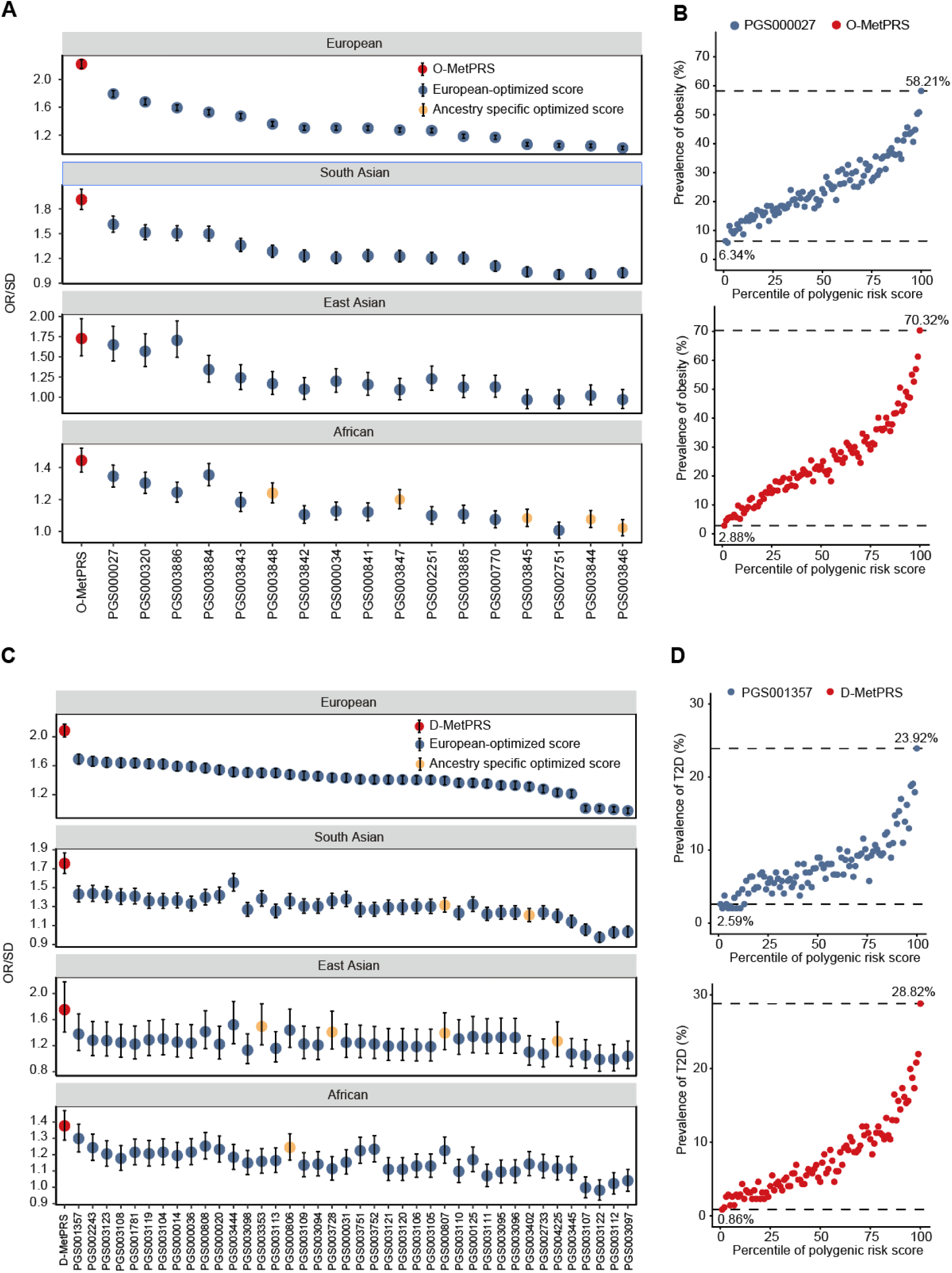
Internal validation of MetPRS performance in UK Biobank hold-out test set. The OR/SD with 95% CI and prevalence (%) across percentile bins were assessed for obesity **(A,B)** and T2D **(C,D)** in UK Biobank hold out test set (n = 49,377). MetPRS was compared with all obesity and T2D PRSs published in PGS Catalog (https://www.pgscatalog.org/) that had not included UK Biobank in score development. We then selected highest-performing score among published studies for obesity (PGS000027 - GPS_BMI - Khera AV et al) and T2D (PGS001357 - T2D_AnnoPred_PRS - Ye et al), and compared them with MetPRS by grouping according to polygenic risk score percentiles. Obesity was defined as a BMI of 25 Kg/m^2^ or above for East Asians, and 30 Kg/m^2^ or above for all other populations. O-MetPRS: obesity MetPRS; D-MetPRS: T2D MetPRS; OR/SD: odds ratio per increase in 1 standard deviation.

We further tested the associations of O-MetPRS and D-MetPRS with various metabolic traits (**Extended Data Fig.2 and Supplementary Table 6**). A consistent significance across all traits (P < 0.0001) suggested that both O-MetPRS and D-MetPRS effectively capture the genetic predispositions related to obesity and T2D, respectively. O-MetPRS demonstrated large association coefficients observed for weight (β = 4.75 Kg per SD, SE = 0.07, P < 0.0001), hip circumference (β = 2.95 cm per SD, SE = 0.047, P < 0.0001), and BMI (β = 1.86 Kg/m^2^ per SD, SE = 0.024, P < 0.0001). D-MetPRS showed strong associations with HbA1c (β = 1.434 mmol/mol per SD, SE = 0.034, P < 0.0001), triglycerides (TG) (β = 6.53 mg/dl per SD, SE = 0.003, P < 0.0001), and glucose (β = 3.098 mg/dl per SD, SE = 0.007, P < 0.0001). Notably, O-MetPRS has a larger effect size on obesity-related traits and D-MetPRS has larger effect size on diabetes-related traits, suggesting that each score captured its relevant biological insights.

### External validation of MetPRS in multiple cohorts

We validated the MetPRS in three external multiethnic cohorts: All of Us (n = 245,394), Mass General Brigham (MGB) Biobank (n = 53,306), and the King Faisal Specialist Hospital and Research Center [KFSH&RC] cohort (n = 6,416) (**Fig. 4; Supplementary Tables 7-12**). MetPRS was compared against the top 5 obesity and T2D PRSs from the PGS Catalog.^13^ We selected the top 5 PRSs that consistently ranked highly across all external datasets as comparators, excluding any scores that incorporated each test cohort (i.e., All of Us) in their score development. The OR/SD with 95% CI for prevalent obesity and T2D was assessed for O-MetPRS and D-MetPRS, respectively, in a logistic regression model adjusted for age, sex, genotyping array, and the first 10 principal components of ancestry. MetPRS was the best-performing score across diverse ancestries, except for O-MetPRS in the East/South Asian population of MGB Biobank, D-MetPRS in Middle Eastern population of All of Us, and D-MetPRS in East Asian population of MGB Biobank (**Fig.4**). In addition to predicting binary obesity, we also evaluated O-MetPRS for its ability to predict continuous BMI (**Extended Data Fig.3**). O-MetPRS outperformed the top five comparator scores across diverse ancestries in predicting BMI, with the exception of East/South Asian and African populations in the MGB Biobank (**Extended Data Fig.3 and Supplementary Table 13**). The consistently high performance across multi-ancestry external cohorts underscored the robustness and generalizability of MetPRS.

**Fig 4.**
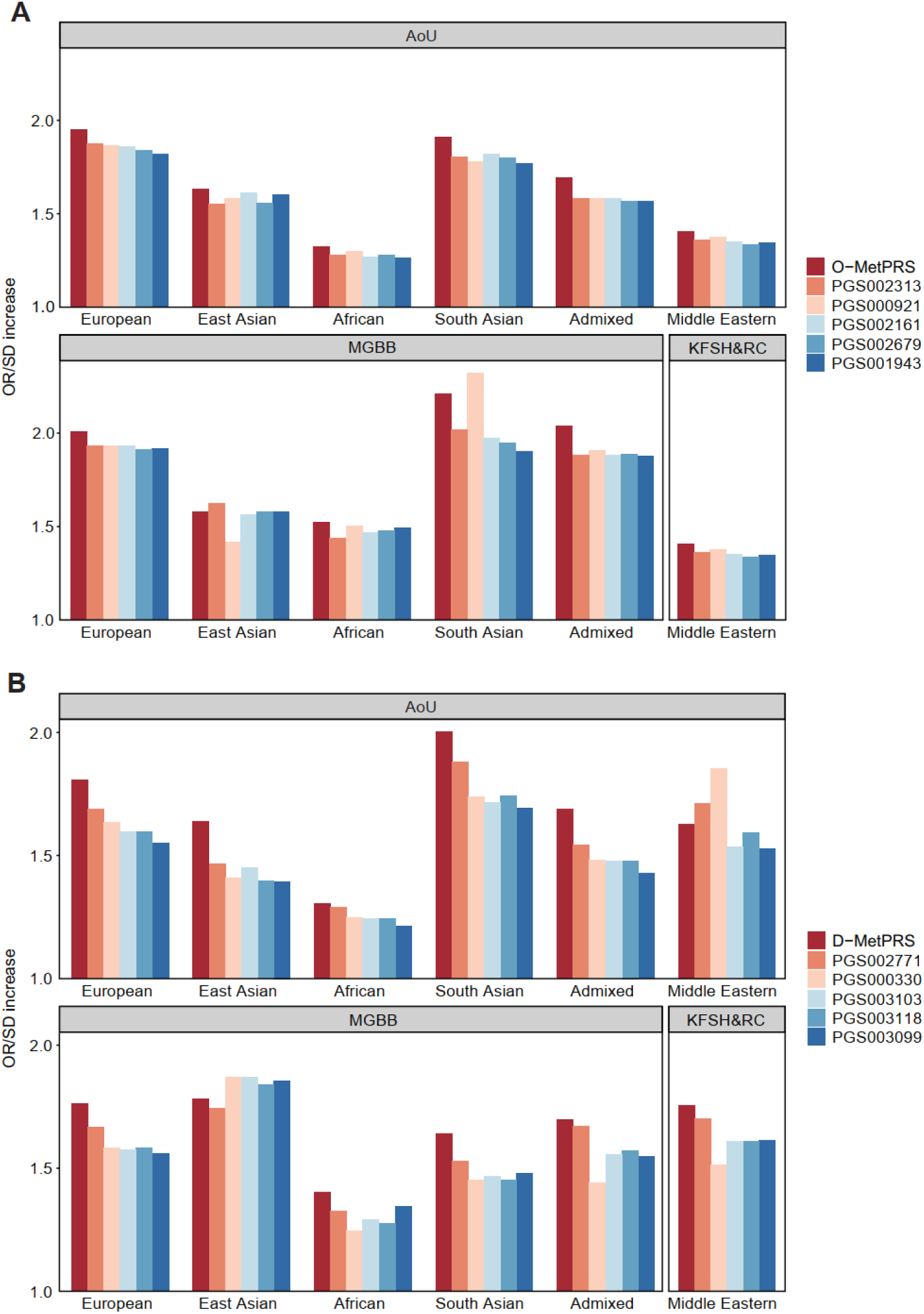
External validation of MetPRS in multiple ancestries. The OR/SD for prevalent obesity **(A)** and T2D **(B)** was assessed for each polygenic risk score in a logistic regression model adjusted for age, sex, genotyping array and the first ten principal components of ancestry in external datasets: All of Us (n = 245,394), MGB Biobank (n = 53,306), and KFSH&RC cohort (n = 6,416). MetPRS was compared with top 5 obesity and T2D scores in PGS Catalog. Polygenic risk scores that consistently ranked highly across external datasets were selected as comparators (top 5), excluding any scores that incorporated any test cohort (i.e., All of Us) in their score development. Obesity was defined as a BMI of 25 Kg/m^2^ or above for East Asians, and 30 Kg/m^2^ or above for all other populations. AoU: All of Us; MGBB: Massachusetts General Brigham Biobank; Admixed: Latino/admixed American; O-MetPRS: obesity MetPRS; D-MetPRS: T2D MetPRS; OR/SD: odds ratio per increase in 1 standard deviation of PRS score.

Although MetPRS consistently performed better than pre-existing scores across ancestries in relative space, its performance was suboptimal in individuals of African ancestry (**Fig. 4**). To address this gap, we developed an African-optimized MetPRS by training the model in UK Biobank African participants, which led to a further performance improvement tested in the All of Us and MGB Biobank (**Extended Data Fig.4**).

### Prediction of Morbidities Related to Obesity and Type 2 Diabetes

Obesity and T2D are not only major health challenges on their own but also serve as strong risk factors and precursors for a wide array of downstream diseases and complications. We assessed the ability of MetPRS to predict morbidities related to obesity and T2D using a Cox proportional hazards model for individuals in the MetPRS top decile compared to those of the rest cohort; the risk of all tested morbidities was significantly higher in top decile group for O-MetPRS and D-MetPRS (**Extended Data fig.5; Supplementary Table 14**). When compared to the top 5 comparator scores, O-MetPRS had the best prediction of obesity-related morbidities, ranking first for predicting 12 out of 14 obesity-related morbidities such as hypertension (hazard ratio[HR] 1.39; 95% CI: 1.30-1.49), venous thromboembolism (HR1.47; 95% CI: 1.21-1.80), and Nonalcoholic fatty liver disease (HR 1.83; 95% CI: 1.41-2.37) (**Fig. 5A and 5B; Extended Data Fig. 6**). Similarly, D-MetPRS most accurately predicted diabetes-related morbidities, ranking first for predicting 11 out of 13 diabetes-related morbidities such as neuropathy (HR 1.87; 95% CI: 1.35-2.59), nephropathy (HR 1.89; 95% CI: 1.65-2.16), and retinopathy (HR 1.87; 95% CI: 1.35-2.59) (**Fig. 5C and 5D; Extended Data Fig. 6**).

**Fig 5.**
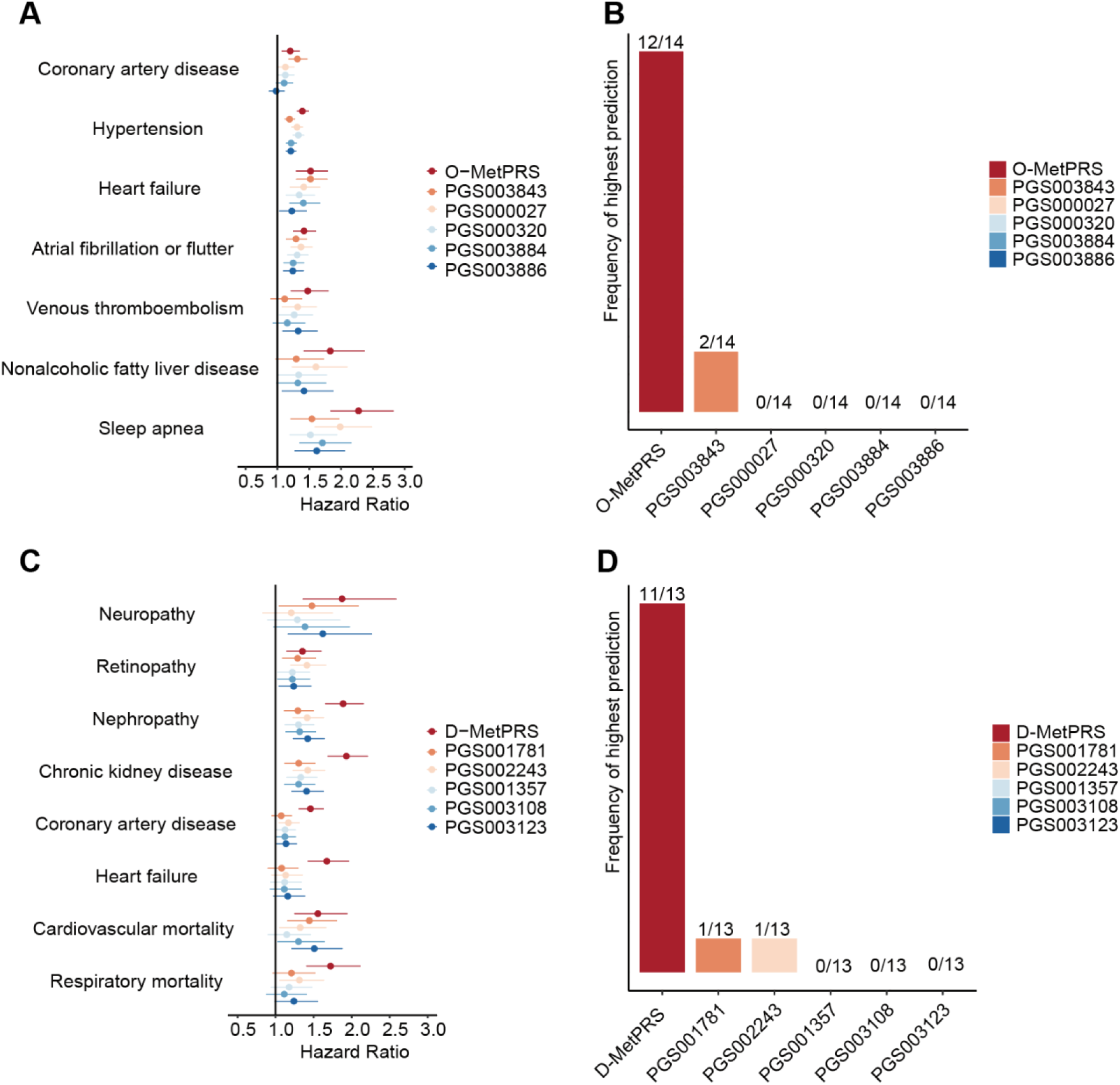
Prediction of obesity and diabetes-related morbidities. The top decile of the MetPRS distribution was identified as "carriers," and their risk was assessed against the rest of the cohort. Risk was estimated using a Cox proportional hazards regression model, adjusted for age, sex, genotyping array and the first ten principal components of ancestry. The performance of O-MetPRS and the other top scores in predicting obesity-related morbidities was evaluated **(A)**, and their corresponding rank frequencies across morbidities were summarized **(B)**. The performance of D-MetPRS and other top scores in predicting diabetes-related morbidities and complications was evaluated **(C),** and their corresponding rank frequencies across morbidities and complications were summarized **(D)**. The forest plot for the other conditions refers to Extended Data Fig.5. Full list of morbidities is provided in Supplementary Table 14.

### MetPRS and Risk of GLP-1 Receptor Agonist Prescription and Metabolic-bariatric Surgery

The MGB Biobank consisted of 51,779 eligible participants, of whom 4,490 (8.7%) were prescribed GLP-1 receptor agonists during a mean follow-up of 5.53 (SD 2.96) years, and 1,487 (2.9%) underwent metabolic-bariatric surgery during a mean follow-up of 5.53 (SD 3.32) years. Among 51,779 participants, 37,329 were free of obesity and T2D at baseline. To evaluate the relationship between MetPRS and clinical interventions in contemporary practice, we employed Cox proportional hazards regression models, adjusting for age, sex, and top 10 ancestry principal components. Participants were categorized into three risk groups based on their MetPRS – the top 10%, middle quintile (40–60%), and bottom 10%.

Among 37,329 MGB individuals free of obesity and T2D, those in the top decile compared to the population median quintile of O-MetPRS had a markedly increased risk of receiving a GLP-1 agonist prescription (HR 2.03; 95% CI: 1.65-2.50) and metabolic-bariatric surgery (HR 3.61; 95% CI: 2.06-6.33). The cumulative incidence of GLP-1 receptor agonist prescriptions and metabolic-bariatric surgery showed significant variation across different O-MetPRS risk groups at 10-year (**Fig. 6A and 6B; Extended Data Fig. 7A and 7B**). Among the group free of obesity and T2D at baseline, participants in the top 10% risk group had a cumulative incidence of 12.6% for GLP-1 receptor agonist prescriptions and 0.4% for metabolic-bariatric surgery. For the middle quintile group, the cumulative incidences were 6.5% and 0.1%, while the bottom 10% group had incidences of 3.4% and 0.0%, respectively (**Fig.6A and 6B**). These trends were consistent in the overall population, which included some participants with obesity and T2D at baseline who had not been treated with these interventions. The cumulative incidence of GLP-1 receptor agonist prescriptions and metabolic-bariatric surgery was generally higher in this population (**Extended Data Fig. 7A and 7B**).

**Fig 6.**
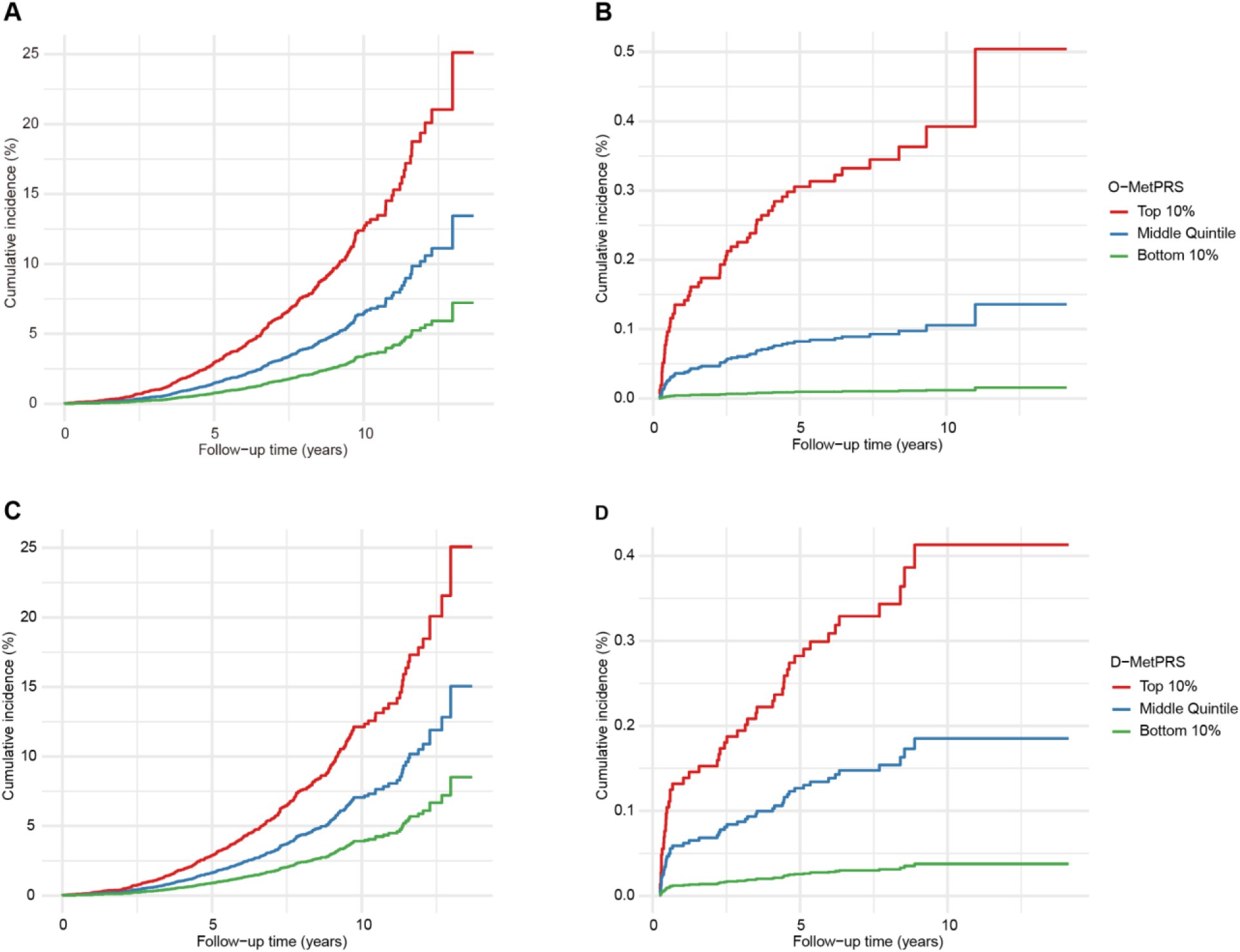
Risk for incident GLP-1 receptor agonist prescription and metabolic-bariatric surgery according to MetPRS strata in MGB Biobank participants free of obesity and T2D at the baseline. The 37,329 participants without baseline obesity and T2D were included for analysis. Cumulative incidence of GLP-1 receptor agonist prescription **(A)** and metabolic-bariatric surgery **(B)** stratified by O-MetPRS; cumulative incidence of GLP-1 receptor agonist prescription **(C)** and metabolic-bariatric surgery **(D)** stratified by D-MetPRS. The risks were estimated using Cox proportional hazards regression model adjusted for age, sex, and the first ten principal components of ancestry in a 37,329 population without obesity and T2D at the baseline in MGB Biobank. GLP-1 receptor agonist prescription and metabolic-bariatric surgery were selected as clinical endpoints as they are interventions indicated for obesity or diabetes or both.

Similarly, those in the top decile compared to the population median quintile of D-MetPRS had a substantially increased risk of receiving a GLP-1 agonist prescription (HR 1.80; 95% CI: 1.47-2.21) and metabolic-bariatric surgery (HR 2.29; 95% CI: 1.29-4.06). The cumulative incidence of GLP-1 receptor agonist prescriptions and metabolic-bariatric surgery also varied notably across the D-MetPRS risk groups at the 10-year mark (**Fig. 6C and 6D; Extended Data Fig. 7**). In the population, those in the top 10% risk category had cumulative incidences of 12.1% for GLP-1 receptor agonist prescriptions and 0.4% for metabolic-bariatric surgery. In contrast, the middle quintile group had cumulative incidences of 7.0% and 0.2%, and the bottom 10% group had 3.9% and 0.0%, respectively (**Fig.6C and 6D**). These patterns persisted in the overall population with higher incidence rates (**Extended Data Fig. 7C and 7D**).

## Discussion

This study introduces MetPRS for diabesity based on a novel approach that captures the genetic architecture of metabolic dysfunction by synthesizing genetic association data from related measures and indices. This method leveraged biological insights to optimize genetic risk prediction for obesity, T2D, their related morbidities, and even clinical interventions in contemporary practice. We demonstrate that MetPRS outperformed all previous PRSs in predicting obesity and T2D across multiple ancestries, including European, African, East Asian, South Asian, Latino/admixed American, and Arab populations. Moreover, MetPRS precisely predicted downstream morbidities and clinical interventions associated with diabesity, such as GLP-1 receptor agonist prescriptions and metabolic-bariatric surgery, underscoring its potential clinical utility. The biologically enriched MetPRS is well-positioned to penetrate various aspects of clinical practice, from predicting morbidities to guiding management decisions.

Metabolic disorders are inherently multifactorial and involve complex biology. As such, integrating GWAS data for various diabesity-related measures resulted in a biologically-enriched PRS. No single metric can fully capture the etiology of metabolic disorders, necessitating the complementary use of multiple measures in clinical practice. For instance, HbA1c and fasting glucose levels offer complementary insights – HbA1c reflects long-term glycemic control, while fasting glucose levels indicate immediate glucose control.^14^ Likewise, multiple obesity measures, such as BMI, WC, WHR, VAT, ASAT, and GFAT provide both converging and diverging insights into different obesity subtypes and risk trajectories for obesity related morbidities.^15^ This growing body of evidence supported our strategy to develop a biologically-enriched PRS by leveraging genetic information across multiple diabesity-related measures. Emdin et al. previously demonstrated that a PRS for WHR adjusted for BMI (WHRadjBMI) could capture a genetic predisposition to adiposity that was not evident when using the genetic risk of WHR or BMI alone.^16^ Meanwhile, their study was limited to combining only two metrics and was not specifically aimed at developing PRS that captures whole dimensions of underlying etiologies. We integrated a wider range of measures (22 measures), creating a metabolic PRS that is more comprehensive and better equipped to reflect the complex and heterogeneous nature of metabolic disease manifested by obesity and T2D.

O-MetPRS and D-MetPRS demonstrated superior predictive accuracy compared to previously developed PRSs for obesity and T2D in the PGS catalog in both internal and external validations (**Fig. 3 and 4**). The genome-wide PRS for obesity (GPS_BMI_) introduced by Khera et al.(PGS000027),^7^ which utilized 2.1 million genetic variants, had previously set the benchmark for obesity PRS performance. However, our O-MetPRS showed a significant improvement, with an over 20% increase in OR/SD compared to GPS_BMI_ (GPS_BMI_ OR/SD 1.79 vs. O-MetPRS OR/SD 2.22 in European population) (**Fig. 3**). This substantial enhancement could be attributed to the difference in genetic inputs and reflecting their biological relevance (**Extended Data Fig.8**); O- MetPRS incorporated multiple ancestry GWAS from 22 traits, whereas GPS_BMI_ relied on only BMI and European GWAS data. Similarly, D-MetPRS demonstrated over 20% increase in OR/SD compared to the second highest T2D PRS––AnnoPred PRS (PGS001357) (AnnoPred OR/SD 1.69 vs. O-MetPRS OR/SD 2.08 in European population).^17^ Such improvement over previous top scores for both obesity and T2D underscores the value of our multiplicative approach.

Obesity and T2D are not only major health concerns in their own right but also strong risk factors for numerous morbidities,^18–22^ making it important to predict their related comorbidities for better health management. Our study demonstrates that MetPRS had a substantial improvement in predicting downstream morbidities and complications of diabesity compared to existing PRSs (**Fig. 5 and Extended Data Fig.6**), likely due to its biologically enriched composition. Moreover, MetPRS effectively predicted cardiovascular mortality, likely indicating its ability to identify individuals at risk for poorer outcomes or prognosis, beyond merely predicting disease incidence. Few AI-based tools and clinical scores have been developed to predict obesity-related or diabetic complications, yet they require clinical factors that usually become evident in middle age as inputs.^23–25^ In contrast, MetPRS can predict morbidities long before the clinical factors manifest, potentially as early as birth.^7^ As sequencing technologies advance and become more cost-effective and affordable,^26,27^ the use of MetPRS can shift morbidity care earlier in life and support primordial prevention.

The MetPRS was able to predict the likelihood of requiring GLP-1 receptor agonist prescription in a population free of obesity and T2D at baseline, over a period of following up to 13 years. The GLP-1 receptor agonists are indicated for treating obesity and T2D, yet their indications are expanding as mounting evidence shows their efficacy in reducing cardiovascular diseases,^28^ steatohepatitis,^29^ chronic kidney disease,^30^ and other conditions.^31,32^ Despite its growing status as a widely used treatment, the challenges in the pharmacological intervention lie in evaluating the risk-benefit ratio for each individual, as GLP-1 receptor agonists are not without adverse effects.^33,34^ An even greater concern is the significant economic burden these medications impose on healthcare systems, particularly in countries like the US, where up to 40% of the population may be eligible for treatment.^35,36^ Therefore, the ability to extend genetic prediction to GLP-1 receptor agonist prescription holds clinical implications, adding a data point for risk-benefit assessment, patient-physician communication, and cost-effective rollout. Currently, GLP-1 receptor agonists are typically reserved for individuals with obesity or diabetes who have comorbidities or have not achieved sufficient control through lifestyle modifications.^37,38^ The ability to genetically predict the eventual need for this drug could potentially move the needle in management plans and health system strategies.

MetPRS is a multi-ancestry PRS that can be readily applied to diverse populations, as it showed robust prediction performance across different ancestries in our external validation data sets (**Fig. 4**). The Eurocentric bias inherent in many genetic studies has significantly limited the generalizability of PRSs, presenting a major barrier to their implementation in real-world settings where populations are inherently diverse.^39–42^ Our study addressed this challenge by incorporating GWAS data from multiple ancestries when developing the model, ensuring that genetic information from non-European populations is represented. We then validated the MetPRS across multiple ancestries, demonstrating consistent best performances across these populations. Notably, even though some existing PRSs were optimized for their specific non-European ancestries, our multi-ancestry MetPRS still outperformed them in respective non-

European populations (**Fig. 3 and Fig. 4**). Given its reliable performance among various ancestries, MetPRS can be readily implemented in ethnically diverse settings and contexts. Our findings contribute to the growing body of evidence that multi-ancestry genetic studies can facilitate more inclusive and widely applicable health services and alleviate the risk of propagating potential healthcare disparities in genomic medicine.^8,43^

There are several limitations that must be acknowledged. First, while we incorporated multi-ancestry and non-European GWAS data to train MetPRS, the majority of the data used for model development and validation is still derived from GWAS of European descent, which may affect the score’s performance in non-European populations. Reassuringly, MetPRS consistently performed better than pre-existing scores across ancestries on a relative scale. However, its performance was suboptimal in individuals of African ancestry, and thus we developed an African- optimized MetPRS, which led to a modest improvement in performance. Second, our study centered on obesity and T2D among metabolic traits. Future research could explore the inclusion of additional metabolic traits or different combinations based on biological commonality. However, there are several justifications for our focus. We specifically targeted obesity and T2D due to their closely related pathophysiology^1,2^ and shared treatment options, including GLP-1 receptor agonists and metabolic-bariatric surgery.^44–46^ Convergence in clinical management was a particularly important consideration, as our aim for MetPRS was to design a PRS that can predict across various layers of clinical aspects. The intersecting foundation in biology, etiology, and clinical workflow pertinent to both conditions enabled MetPRS to be a clinically relevant PRS. Finally, the clinical implementation of MetPRS requires further consideration of how to interpret and communicate PRS results to healthcare providers and patients, as well as how to measure its impact on patient outcomes and healthcare decision-making.^47^

## Conclusion

This study presented a novel approach to genetic risk prediction of diabesity by introducing the MetPRS, a composite metabolic PRS that integrates multi-ancestry, multi-disease, and multi-measure genetic data. Our findings have significant implications for precision medicine, offering more comprehensive scores to predict risks for obesity and T2D, as well as their complications and clinical management. The MetPRS outperformed traditional single-trait PRS models, demonstrating the value of a biologically-enriched approach to genetic risk prediction. By leveraging multi-ancestry GWAS data, MetPRS represents a step forward in creating more equitable and inclusive tools for genomic medicine.

## Methods

### Study cohorts

#### UK Biobank

The UK Biobank (UKB) is an observational study that enrolled over 500,000 individuals aged 40 to 69 across the United Kingdom between 2006 and 2010. At the time of recruitment, participants provided electronically-signed consent, completed questionnaires on socio- demographic, lifestyle, and health-related factors, and underwent various physical assessments.^48^ Anthropometric measurements, including body mass index, were measured at the initial enrollment visit. Among participants, 43,521 underwent MRI imaging between 2014 and 2020.^49,50^ Participants were genotyped using the UK BiLEVE Axiom Array or the UK Biobank Axiom Array, each containing over 800,000 genome-wide variants. Imputation was carried out using reference panel from the Haplotype Reference Consortium (HRC),^51^ UK10K,^52^ and the 1000 Genome Phase 3 data.^53^ The HRC was used as the primary imputation reference panel due to its large sample size (64,976 broadly European haplotypes). The 1000 Genomes phase 3 dataset was employed to assist with the phasing of samples from non-European ancestries. Imputation results were also combined from the merged UK10K and 1000 Genomes phase 3 reference panels and the HRC panel, with HRC imputation being prioritized when a SNP was present in both panels. Imputation was conducted using IMPUTE 4 (https://jmarchini.org/software/).

#### All of Us

The All of Us (AoU) Research Program is a longitudinal cohort study that has enrolled a diverse group of traditionally underrepresented individuals aged 18 and older from more than 730 sites across the United States.^54^ Since its inception in 2018, the AoU program has consented to over 800,000 participants, with more than 560,000 completing the initial enrollment process, including health questionnaires and biospecimen collection. For these participants, ongoing linkage to electronic health record (EHR) data, including International Classification of Diseases (ICD)-9/ICD-10, Systematized Nomenclature of Medicine (SNOMED), and Current Procedural Terminology (CPT) codes, is maintained. This study utilized whole-genome sequencing (WGS) data from over 245,000 participants available in the Controlled Tier Dataset, version 7 release. Data analysis was conducted on the All of Us Researcher Workbench under the guidelines defined by the All of Us Ethical Conduct of Research Policy.

#### Mass General Brigham Biobank

The Mass General Brigham Biobank (MGBB) is a large-scale, healthcare-affiliated biobank based in the Greater Boston area of Massachusetts.^55,56^ Established in 2010, the MGBB has enrolled approximately 140,000 diverse individuals within the Mass General Brigham (MGBB) network, the largest healthcare system in Massachusetts. The biobank aims to uncover the complex relationships among genomic profiles, environmental factors, and disease manifestations in clinical practice. MGBB provides biomedical samples, such as plasma, serum, DNA, buffy coats, collected from patients and linked to clinical data from the EHR, quantitative data derived from medical images, and survey data on lifestyle, environment, and family history.^57^ Genome-wide genotyping arrays of 53,306 MGBB participants were used in this study.

#### King Faisal Specialist Hospital and Research Center cohort

The King Faisal Specialist Hospital and Research Center (KFSH&RC) cohort consists of indigenous Arabs from Saudi Arabia referred for cardiology care at KFSH&RC, a tertiary care hospital recognized throughout Saudi Arabia and the Middle East, for work-up of cardiac disease from all over the country^58^. In addition to blood samples for DNA extraction and genotyping array analyses, participants provided access to their electronic health records for phenotype data and shared socio-demographic information through a clinical research coordinator. All participants gave informed consent to participate in the study.

### Phenotype definitions

For the UK Biobank participants, weight and height were collected at baseline when participants attended the initial assessment center. BMI was constructed from height and weight measured during the initial Assessment Centre visit. BMI is not present if either of these readings were omitted. Obesity was defined as a BMI ≥ 25 kg/m² for individuals of East Asian ancestry and a BMI ≥ 30 kg/m² for individuals of other ancestries,^59,60^ which was used also for AoU, MGBB and KFSH&RC cohorts. T2D was identified either through self-reported diagnosis during an interview with a trained nurse or via the ICD-10 code E11.X in hospitalization records. For comorbidities, detailed definitions of diseases included in the comorbidities analysis are provided in **Supplementary Table 17**. For example, coronary artery disease cases were defined centrally based on self-report at enrollment, hospitalization records, or death registry records; retinopathy case ascertainment was based on hospitalization records and death registry records; sleep apnea ascertainment was based on self-report at the time of enrollment, hospitalization records, and death registry records.

For All of Us, BMI was taken from physical measurements of EHR data. T2D was identified as EHR records containing the term “Type 2 diabetes mellitus” in Controlled Tier Dataset v7 Conditions. For MGBB, BMI was obtained from the electronic health records T2D was identified as glycated hemoglobin ≥ 6.5% or diagnosis records from the electronic health records. For the KFSH&RC cohort, disease status and metabolic traits were collected during routine clinical care and manually extracted from the electronic health record by trained medical personnel. The diagnosis of T2D was curated from the electronic health records. Height and weight measurements obtained in the context of care and the first visit measured on referral to the King Faisal Specialist Hospital and Research Center was used.

### Define polygenic risk score percentiles

Residual scores for MetPRS, and PRS scores calculated by the variant weight from the PGS Catalog were extracted after adjustment for the first ten principal components of genetic ancestry, then normalized to a mean of 0 with a standard deviation of 1. The percentile distribution of the polygenic risk scores was analyzed to assess the stratification of obesity (BMI ≥ 30 kg/m²) and T2D.

### Quality control and sample selection

#### UK Biobank

Participants were excluded from the analysis if they met any of the following criteria: (1) a mismatch between self-reported sex and genetically inferred sex, (2) sex chromosome aneuploidy, (3) a genotyping call rate below 0.95, (4) being outliers for heterozygosity, (5) having second-degree or closer relatives, (6) with unknown self-reported ancestry, or (7) without BMI or T2D phenotypes. After applying these sample quality controls, 444,345 participants remained available (**Extended Data Fig.1**).

To further boost the power for constructing PRS scores, we conducted de novo GWAS for ten traits (BMI, T2D, body fat percentage, height, body weight, waist circumference, hip circumference, HbA1c, Glucose, cT1) traits utilizing data from 378,921 participants randomly selected from UKB European ancestry participants (self-reported ancestry, Field ID 21000). For the remaining participants of European ancestry, 15,000 and 34,626 individuals were assigned to the training and testing sets for PRS score development, respectively. A total of 1,304 individuals who self-identified as Chinese were included for East Asian ancestry validation, 6,842 individuals who self-identified as Black were included for African ancestry validation, and 6,605 individuals of South Asian descent (self-reported as Indian, Pakistani, or Bangladeshi) were included for South Asian ancestry validation. Individuals with body (abdominal) magnetic resonance imaging (MRI) data (UK Biobank Field ID 12224) were excluded from the training and validation datasets, since GWAS for MRI-derived visceral (VAT), abdominal subcutaneous (ASAT), and gluteofemoral (GFAT) adipose tissue volumes was conducted on UKB MRI-tested participants;^61^ we additionally excluded individuals with whole body DXA scan (UK Biobank Field ID 20201), (**Extended Data Fig.1**).

Genetic variants were included in the GWAS if they met the following quality control criteria: (1) imputation quality score (INFO) > 0.3, (2) minor allele frequency (MAF) > 0.001, (3) minor allele count (MAC) > 100, and (4) SNP genotyping missing rate < 10%. These criteria resulted in a total of 15,848,715 imputed variants available for analysis. Common variant association studies for ten traits (BMI, T2D, body fat percentage, height, body weight, waist circumference, hip circumference, HbA1c, Glucose, cT1) were conducted using REGENIE (v3.2.7), genotype variants were centrally imputed from UK Biobank.^48^

#### All of Us

Within All of Us, details of whole genome sequencing and quality control are described extensively in Jurgens et al.^62^ and the All of Us Genomic Research Data Quality Report C2022Q4R9 CDR v7, available at https://support.researchallofus.org/hc/en-us/articles/4617899955092-All-of-Us-Genomic-Quality-Report. Briefly, 229,517 out of total 245,394 samples remained after filtering for sex concordance, a cross individual contamination rate below 3%, a call rate above 98% and related samples by AoU Central quality control. We used the Allele Count/Allele Frequency (ACAF) threshold SNP callset curated by AoU, which includes SNPs of population-specific allele frequency > 1% or allele counts over 100 for each ancestral subpopulation. The inferred ancestry information is obtained from the “ancestry_pred” column, which is available as part of the genetically predicted ancestry TSV file. The quality control metrics were centrally provided by the AoU project.^54^ SNPs deviated from Hardy-Weinberg equilibrium (P < 1×10^−6^) in each genetic ancestry were removed. Principal components of genetic ancestry used for the correction of population structure were calculated in each ancestry group separately using SNPs present in the 1000 Genomes project phase 3 release.

#### Mass General Brigham Biobank

Within MGB Biobank, 53,306 individuals were genotyped by Illumina Global Screening Array (Illumina, CA) and imputed to the TOPMed multi-ancestry imputation reference panel (TOPMed r2 panel).^56^ Variants with high missingness (> 2%) and low MAF (< 1%) and variants that failed the Hardy–Weinberg test (*P* < 1 × 10^−6^) were removed. After excluding individuals due to discrepancies between reported and genotypic sex, sex chromosome aneuploidy, genotyping call rate < 0.95, heterozygosity outliers, and excess relatedness (second-degree relatives or closer), 49,825 individuals remained for validation. We inferred genetic ancestry using a public diverse population, including 3,380 unrelated individuals from the 1000 Genomes Project (1KG) and the Human Genome Diversity Project (HGDP), called 1KG_HGDP.^63^ We extracted common, high-quality SNPs (missingness < 1%, MAF > 1%) across MGBB and the 1KG_HGDP dataset. After pruning SNPs, we computed SNP weights for the genetic principal component using the 1KG_HGDP dataset. Then, we projected MGBB participants into the same principal component space using the top 20 PCs. Using genetic PCs in the 1KG_HGDP dataset as a feature matrix, we trained a K-nearest neighbor model (k=1) for 1KG_HGDP reference populations to assign population labels to MGBB participants.

#### King Faisal Specialist Hospital and Research Center cohort

All participants from the King Faisal Specialist Hospital and Research Center cohort were of Middle Eastern ancestry and variants and sample quality control were previously described.^58^ Briefly, we excluded samples due to variant calling missingness > 5%, heterozygosity rate > 5 standard deviations above the mean, and mismatch between genotypically-determined and self-reported sex. Variant-level quality control was conducted to remove variants with call rate < 98%, MAF < 0.01, or Hardy–Weinberg test (*P* < 1 × 10^−6^). After imputation by TOPMed r2 panel datasets, the variants with INFO < 0.3 and MAF < 0.01 were removed.

### Genetic correlation estimation between traits

We selected 22 traits that directly or indirectly serve as proxies for the status of obesity and T2D (**Supplementary Table 1**). Our focus on obesity and T2D among various metabolic diseases is due to their closely related pathophysiology and shared treatment options, including GLP-1 receptor agonists and metabolic-bariatric surgery. This common biological, etiological, and clinical foundation justified the development of a composite metabolic PRS for diabesity. We defined a ‘trait’ as a composite of binary disease status (e.g., T2D) and continuous measures for metabolic status (e.g., BMI, HbA1c).

Genetic correlations between 22 traits were estimated using cross-trait LD-score regression (LDSC v1.0.1). Precomputed LD scores were obtained from ∼1.2 million common SNVs in the well-imputed HapMap3 variants panel.^64^ For each trait, only GWAS with the largest sample size of European individuals was used to perform cross-trait LD-score regression analysis. A Bonferroni corrected P value threshold of 0.00227 (0.05/22) was used to define statistical significance.

### Polygenic risk score derivation

Summary statistics from recent obesity and T2D-related GWAS studies (**Supplementary Table 1**) conducted in diverse ancestries were used to determine the primary weights for obesity and T2D PRSs. To ensure an independent holdout dataset for training and validating the obesity and T2D metabolic polygenic risk scores (O-MetPRS and D-MetPRS), UK Biobank participants for de novo GWAS were excluded for PRS score development (**Extended Data Fig.1**). Ancestry-specific linkage disequilibrium (LD) reference panels were derived from the 1000 Genomes Project phase 3 data to match the ancestry of the discovery GWAS.

The construction of O-MetPRS and D-MetPRS involved a two-layer process (**Fig.1B**). In layer 1, multiple PRS derived from ancestry-specific GWAS data were combined to yield a multi-ancestry PRS for each trait. In layer 2, the multi-ancestry PRSs for 22 traits were further combined to generate MetPRS.

In layer 1, separate PRSs were constructed based on each ancestry-stratified GWAS using the LDpred2 method, a Bayesian approach that calculates posterior mean effects for all variants, considering prior GWAS effect sizes and subsequent shrinkage based on LD.^65^ Only HapMap3 variants—a set of 1,296,172 variants representing common genetic variation across diverse populations—were included in the score calculation.

The LDpred2 method used default parameters, including proportion of causal variant (values of P = 1.0 × 10^−4^, 1.8 × 10^−4^, 3.2 × 10^−4^, 5.6 × 10^−4^, 1.0 × 10^−3^, 1.8 × 10^−3^, 3.2 × 10^−3^, 5.6 × 10^−3^, 1.0 × 10^−2^, 1.8 × 10^−2^, 3.2 × 10^−2^, 5.6 × 10^−2^, 1.0 × 10^−1^, 1.8 × 10^−1^, 3.2 × 10^−1^, 5.6 × 10^−1^ and 1), heritability scales (s = 0.7, 1, and 1.4), and whether a sparse LD matrix was applied. These parameter combinations yielded 102 candidate PRSs for each GWAS summary statistic. Genotypes were extracted from a centrally imputed data repository by the UK Biobank, processed using bgenix,^66^ and PRSs were calculated for each individual in the UK Biobank using PLINK 2.0.^67^ The best PRS was selected based on its performance in predicting obesity or T2D in an independent sample of 15,000 White British individuals from UK Biobank. The obesity model used log(BMI) as the outcome, while the diabetes model used T2D. The best score for each ancestry was selected based on predictive performance measured by odds ratio (binary traits) or incremental R^2^(continuous traits). The discriminative capacities (AIC) were then used to determine the optimal combination of different ancestry scores. A regression model estimated the mixing weights for ancestry-specific PRSs, which were then linearly combined into a single multi-ancestry score. Similar procedures were followed for other diabesity-related traits.

In layer 2, these multi-ancestry trait-specific PRSs were linearly combined with the multi-ancestry obesity/T2D PRSs from layer 1 to generate the final O-MetPRS and D-MetPRS. Same as the layer 1, stepAIC was used for the feature selection for predicting log(BMI) or T2D to identify the best combination of trait-level scores for mixing. Then, a regression model was used to estimate the mixing weights for each individual trait-specific PRS as described above, which were then linearly combined into a single MetPRS (O-MetPRS or D-MetPRS). Among the 93 ancestry- and trait-specific scores analyzed in the MetPRS training, 31 and 37 scores significantly contributed to the overall prediction in O-MetPRS and D-MetPRS, respectively, after optimization with stepAIC and weighting via regressions across both layers. The final mixing weights for each layer were summarized in **Extended Data fig 8**, and were listed in **Supplementary Table 15 and 16**.

### Polygenic risk score validation

MetPRS underwent internal hold-out validation within UK Biobank. To evaluate its performance, we compared it with previously published polygenic risk scores for BMI and T2D from the PGS Catalog (https://www.pgscatalog.org/). The variant effect sizes for these scores were obtained from the PGS Catalog and calculated within the same UK Biobank testing dataset, which included 34,626 European ancestry samples, 6,842 of African ancestry, 1,304 of East Asian ancestry, and 6,605 South Asian ancestry samples (**Extended Data Fig.1**). External validation was also performed in the All of Us (n=245,394), MGB Biobank (n=53,306), and a KFSH&RC cohort (n=6,416).

### GLP-1 agonist prescription and metabolic-bariatric surgery

We conducted a comprehensive analysis using data from the MGB Biobank to evaluate the predictive utility of MetPRS for incident GLP-1 receptor agonist prescriptions and metabolic-bariatric surgeries. GLP-1 receptor agonist prescription and metabolic-bariatric surgery were selected as clinical endpoints as they are interventions indicated for obesity and/or T2D. Initial datasets were filtered to include only individuals with complete genotypic and phenotypic data, excluding those censored before the biobank enrollment date, leaving 51,779 participants for analysis. For each participant, O-MetPRS and D-MetPRS were calculated using ancestry-adjusted models. The first ten principal components were regressed out of the raw O-MetPRS and D-MetPRS using linear regression to account for population stratification. The residuals from this regression, which represent the MetPRS adjusted for ancestry, were then standardized to have a mean of 0 and a standard deviation of 1.

To assess the association of MetPRS with clinical interventions, we utilized Cox proportional hazards regression models adjusted for age, sex, and ancestry PCs. We stratified participants into three risk groups based on their MetPRS values: top 10%, middle quintile (40– 60%), and bottom 10%. The primary outcomes were the cumulative incidence of GLP-1 receptor agonist prescriptions and metabolic-bariatric surgery over the follow-up period. Incidence was tracked from baseline until the first event or censoring. Competing risk models were applied to calculate cumulative incidence functions, and Kaplan-Meier curves were generated to visualize time-to-event data.

### Statistical analysis

For score training and validations, prediction for a continuous trait was calculated using linear regression models, while logistic regression models were employed for predicting the risk of a binary trait. All the models included baseline covariates: enrollment age, sex, genotyping array, and the first ten principal components of genetic ancestry. For the comorbidities analysis, the top decile of the PRS distribution was labeled as "carriers," while the remaining individuals were labeled as "non-carriers". The top five obesity and T2D scores in the score comparisons among 34,626 European individuals from the UK Biobank were compared with the O-MetPRS and D-MetPRS, respectively. Cox proportional-hazards models were used to estimate hazard ratios (HRs) for incident diseases in the UK Biobank, with enrollment age, genetically inferred sex, genotyping array, and the first ten principal components as covariates. The frequency of the highest prediction for each score was then calculated. All statistical analyses were performed using R software (version 4.2.2, R Project for Statistical Computing), and figures were generated using the ggplot2 R package (version 3.4.2).

## Data availability

All data are made available from the UK Biobank (https://www.ukbiobank.ac.uk/enable-your-research/apply-for-access) to researchers from universities and other institutions with genuine research inquiries following institutional review board and UK Biobank approval. This research was conducted using the UK Biobank resource under application number 89885. All of Us data are made available from the *All of Us* Research Study to researchers from universities and other institutions with genuine research inquiries following institutional review board and *All of Us* approval. KFSH&RC and MGBB data are governed by local laws, so relevant data could be made available by contacting the investigators. The genome-wide association data supporting the findings of this study are publicly available in KoGES (https://koges.leelabsg.org/), Biobank Japan (http://jenger.riken.jp/en/result), FinnGen (https://www.finngen.fi/en/access_results), AGEN T2D (https://kp4cd.org/index.php/node/309), GIANT (https://portals.broadinstitute.org/collaboration/giant/) and Million Veteran Program (via dbGaP at https://ftp.ncbi.nlm.nih.gov/dbgap/studies/, under accession number phs001672). The research was approved by the Beijing Institute of Genomics (Chinese Academy of Sciences) and the China National Center for Bioinformation institutional review board, and the Mass General Brigham institutional review board.

## Data Availability

All data are made available from the UK Biobank (https://www. ukbiobank.ac.uk/enable-your-research/apply-for-access) to researchers from universities and other institutions with genuine research inquiries following institutional review board and UK Biobank approval. This research was conducted using the UK Biobank resource under application number 89885. All of Us data are made available from the All of Us Research Study to researchers from universities and other institutions with genuine research inquiries following institutional review board and All of Us approval. KFSH&RC and MGBB data are governed by local laws, so relevant data could be made available by contacting the investigators. The genome-wide association data supporting the findings of this study are publicly available in KoGES (https://koges.leelabsg.org/), Biobank Japan (http://jenger.riken.jp/en/result), FinnGen (https://www.finngen.fi/en/access_results), AGEN T2D (https://kp4cd.org/index.php/node/309), GIANT (https://portals.broadinstitute.org/collaboration/giant/) and Million Veteran Program (via dbGaP at https://ftp.ncbi.nlm.nih.gov/dbgap/studies/, under accession number phs001672). The research was approved by the Beijing Institute of Genomics (Chinese Academy of Sciences) and the China National Center for Bioinformation institutional review board, and the Mass General Brigham institutional review board.

## Acknowledgments

P.N. is supported by grants from the National Institutes of Health (R01HL127564, U01HG011719), P.T.E. is supported by grants from the National Institutes of Health (1R01HL092577, 1R01HL157635, 5R01HL139731), from the American Heart Association Strategically Focused Research Networks (18SFRN34110082), and from the European Union (MAESTRIA 965286). P.N. is supported by grant from National Human Genome Research Institute (U01HG011719). A.C.F. is supported by grants from the National Heart Lung and Blood Institute (K08HL161448 and R01HL164629). Dr. Wang is supported by the Pioneering Action Grants of the Chinese Academy of Sciences. We gratefully acknowledge all participants of UKB, AoU, MGBB and KFSH&RC for their contributions, without whom this research would not have been possible. We also thank the National Institutes of Health’s AoU Research Program, the UKB resource, the MGB team and KFSH&RC team, for making available the participant data examined in this study. Further, we acknowledge the dedication and expertise of the research coordinators and the comprehensive Biobank team, whose efforts in participant enrollment and data collection were indispensable.

## Author contributions

Concept and design: M.S.K, Q.C., P.T.E., A.C.F. and M.W.. Acquisition, analysis or interpretation of data: M.S.K, Q.C., Y.S., X.Y., S.W., L.C.W., S.M.J.C., S.K., X.Y., K.Y., X.C., R.Z., W.Y., S.L., Z.L., A.C.F. and M.W. Drafting of the manuscript: M.S.K, Q.C., Y.S., A.C.F. and M.W. Critical revision of the manuscript for important content: Z.L., F.A., P.N.

## References

1. Ng, A. C., Delgado, V., Borlaug, B. A. & Bax, J. J. Diabesity: the combined burden of obesity and diabetes on heart disease and the role of imaging. Nat. Rev. Cardiol. 18, 291–304 (2021).

2. Chadt, A., Scherneck, S., Joost, H.-G. & Al-Hasani, H. Molecular links between obesity and diabetes:“diabesity”. (2015).

3. Ingelsson, E. & McCarthy, M. I. Human genetics of obesity and type 2 diabetes mellitus: past, present, and future. Circ. Genomic Precis. Med. 11, e002090 (2018).

4. Coral, D. E. et al. A phenome-wide comparative analysis of genetic discordance between obesity and type 2 diabetes. Nat. Metab. 5, 237–247 (2023).

5. Fahed, A. C., Philippakis, A. A. & Khera, A. V. The potential of polygenic scores to improve cost and efficiency of clinical trials. Nat. Commun. 13, 2922 (2022).

6. Hao, L. et al. Development of a clinical polygenic risk score assay and reporting workflow. Nat. Med. 28, 1006–1013 (2022).

7. Khera, A. V. et al. Polygenic prediction of weight and obesity trajectories from birth to adulthood. Cell 177, 587–596. e9 (2019).

8. Patel, A. P. et al. A multi-ancestry polygenic risk score improves risk prediction for coronary artery disease. Nat. Med. 29, 1793–1803 (2023).

9. Maier, R. M. et al. Improving genetic prediction by leveraging genetic correlations among human diseases and traits. Nat. Commun. 9, 989 (2018).

10. Abraham, G. et al. Genomic risk score offers predictive performance comparable to clinical risk factors for ischaemic stroke. Nat. Commun. 10, 5819 (2019).

11. Ross, R. et al. Waist circumference as a vital sign in clinical practice: a Consensus Statement from the IAS and ICCR Working Group on Visceral Obesity. Nat. Rev. Endocrinol. 16, 177–189 (2020).

12. Williamson, A. et al. Genome-wide association study and functional characterization identifies candidate genes for insulin-stimulated glucose uptake. Nat. Genet. 55, 973–983 (2023).

13. Lambert, S. A. et al. The Polygenic Score Catalog as an open database for reproducibility and systematic evaluation. Nat. Genet. 53, 420–425 (2021).

14. Committee, A. D. A. P. P. & Committee, A. D. A. P. P. 2. Classification and diagnosis of diabetes: Standards of Medical Care in Diabetes—2022. Diabetes Care 45, S17–S38 (2022).

15. Lotta, L. A. et al. Association of genetic variants related to gluteofemoral vs abdominal fat distribution with type 2 diabetes, coronary disease, and cardiovascular risk factors. Jama 320, 2553–2563 (2018).

16. Emdin, C. A. et al. Genetic association of waist-to-hip ratio with cardiometabolic traits, type 2 diabetes, and coronary heart disease. Jama 317, 626–634 (2017).

17. Ye, Y. et al. Interactions between enhanced polygenic risk scores and lifestyle for cardiovascular disease, diabetes, and lipid levels. Circ. Genomic Precis. Med. 14, e003128 (2021).

18. Kim, M. S. et al. Causal effect of adiposity on the risk of 19 gastrointestinal diseases: a Mendelian randomization study. Obesity 31, 1436–1444 (2023).

19. Kim, M. S. et al. Association between adiposity and cardiovascular outcomes: an umbrella review and meta-analysis of observational and Mendelian randomization studies. Eur. Heart J. 42, 3388–3403 (2021).

20. Kim, M. S. et al. Association of genetic risk, lifestyle, and their interaction with obesity and obesity-related morbidities. Cell Metab. 36, 1494–1503. e3 (2024).

21. Kim, M. S. et al. Integration of observational and causal evidence for the association between adiposity and 17 gastrointestinal outcomes: An umbrella review and meta-analysis. Obes. Rev. e13823 (2024).

22. Tsilidis, K. K., Kasimis, J. C., Lopez, D. S., Ntzani, E. E. & Ioannidis, J. P. Type 2 diabetes and cancer: umbrella review of meta-analyses of observational studies. Bmj 350, (2015).

23. Al-Sari, N. et al. Precision diagnostic approach to predict 5-year risk for microvascular complications in type 1 diabetes. EBioMedicine 80, (2022).

24. Aminian, A. et al. Predicting 10-year risk of end-organ complications of type 2 diabetes with and without metabolic surgery: a machine learning approach. Diabetes Care 43, 852–859 (2020).

25. Turchin, A., et al. EXIST: EXamining rIsk of excesS adiposiTy—Machine learning to predict obesity-related complications. Obes. Sci. Pract. 10, e707 (2024).

26. Satam, H. et al. Next-generation sequencing technology: current trends and advancements. Biology 12, 997 (2023).

27. Kullo, I. J. et al. Polygenic scores in biomedical research. Nat. Rev. Genet. 23, 524–532 (2022).

28. Lincoff, A. M. et al. Semaglutide and cardiovascular outcomes in obesity without diabetes. N. Engl. J. Med. 389, 2221–2232 (2023).

29. Sanyal, A. J. et al. A Phase 2 Randomized Trial of Survodutide in MASH and Fibrosis. N. Engl. J. Med. 391, 311–319 (2024).

30. Perkovic, V. et al. Effects of semaglutide on chronic kidney disease in patients with type 2 diabetes. N. Engl. J. Med. 391, 109–121 (2024).

31. De Giorgi, R., et al. 12-month neurological and psychiatric outcomes of semaglutide use for type 2 diabetes: a propensity-score matched cohort study. EClinicalMedicine 74, (2024).

32. Kosiborod, M. N. et al. Semaglutide in patients with heart failure with preserved ejection fraction and obesity. N. Engl. J. Med. 389, 1069–1084 (2023).

33. He, L. et al. Association of glucagon-like peptide-1 receptor agonist use with risk of gallbladder and biliary diseases: a systematic review and meta-analysis of randomized clinical trials. JAMA Intern. Med. 182, 513–519 (2022).

34. Sodhi, M., Rezaeianzadeh, R., Kezouh, A. & Etminan, M. Risk of gastrointestinal adverse events associated with glucagon-like peptide-1 receptor agonists for weight loss. Jama 330, 1795–1797 (2023).

35. Baig, K., Dusetzina, S. B., Kim, D. D. & Leech, A. A. Medicare Part D Coverage of Antiobesity Medications — Challenges and Uncertainty Ahead. N. Engl. J. Med. 388, 961– 963 (2023).

36. Wright, D. R., Guo, J. & Hernandez, I. A Prescription for Achieving Equitable Access to Antiobesity Medications. JAMA Health Forum 4, e230493 (2023).

37. Committee, A. D. A. P. P. & Committee, A. D. A. P. P. 9. Pharmacologic approaches to glycemic treatment: Standards of Medical Care in Diabetes—2022. Diabetes Care **45**, S125– S143 (2022).

38. Melson, E., Ashraf, U., Papamargaritis, D. & Davies, M. J. What is the pipeline for future medications for obesity? Int. J. Obes. 1–19 (2024).

39. Kachuri, L. et al. Principles and methods for transferring polygenic risk scores across global populations. Nat. Rev. Genet. 25, 8–25 (2024).

40. Ruan, Y. et al. Improving polygenic prediction in ancestrally diverse populations. Nat. Genet. 54, 573–580 (2022).

41. Ding, Y. et al. Polygenic scoring accuracy varies across the genetic ancestry continuum. Nature 618, 774–781 (2023).

42. Lennon, N. J. et al. Selection, optimization and validation of ten chronic disease polygenic risk scores for clinical implementation in diverse US populations. Nat. Med. 30, 480–487 (2024).

43. Koyama, S. et al. Population-specific and trans-ancestry genome-wide analyses identify distinct and shared genetic risk loci for coronary artery disease. Nat. Genet. 52, 1169–1177 (2020).

44. Kim, M. S. et al. Association of bariatric surgery with indicated and unintended outcomes: An umbrella review and meta-analysis for risk–benefit assessment. Obes. Rev. 25, e13670 (2024).

45. Mingrone, G. et al. Bariatric–metabolic surgery versus conventional medical treatment in obese patients with type 2 diabetes: 5 year follow-up of an open-label, single-centre, randomised controlled trial. The Lancet 386, 964–973 (2015).

46. Brown, E., Heerspink, H. J., Cuthbertson, D. J. & Wilding, J. P. SGLT2 inhibitors and GLP-1 receptor agonists: established and emerging indications. The Lancet 398, 262–276 (2021).

47. Vogan, K. Implementing polygenic risk scores in the clinic. Nat. Genet. 56, 557–557 (2024).

48. Bycroft, C. et al. The UK Biobank resource with deep phenotyping and genomic data. Nature 562, 203–209 (2018).

49. Littlejohns, T. J. et al. The UK Biobank imaging enhancement of 100,000 participants: rationale, data collection, management and future directions. Nat. Commun. 11, 2624 (2020).

50. Sudlow, C. et al. UK biobank: an open access resource for identifying the causes of a wide range of complex diseases of middle and old age. PLoS Med. 12, e1001779 (2015).

51. the Haplotype Reference Consortium. A reference panel of 64,976 haplotypes for genotype imputation. Nat. Genet. 48, 1279–1283 (2016).

52. The UK10K Consortium et al. The UK10K project identifies rare variants in health and disease. Nature 526, 82–90 (2015).

53. The 1000 Genomes Project Consortium et al. A global reference for human genetic variation. Nature 526, 68–74 (2015).

54. All of Us Research Program Genomics Investigators. Genomic data in the All of Us Research Program. Nature, 627(8003):340-346 (2024)

55. Boutin, N. T. et al. The evolution of a large biobank at Mass General Brigham. J. Pers. Med. 12, 1323 (2022).

56. Koyama, S. et al. Decoding Genetics, Ancestry, and Geospatial Context for Precision Health. medRxiv 2023.10.24.23297096 (2023).

57. Bhattacharya, R. et al. Clonal Hematopoiesis Is Associated With Higher Risk of Stroke. Stroke 53, 788–797 (2022).

58. Shim, I. et al. Clinical utility of polygenic scores for cardiometabolic disease in Arabs. Nat. Commun. 14, 6535 (2023).

59. Haam, J.-H. et al. Diagnosis of Obesity: 2022 Update of Clinical Practice Guidelines for Obesity by the Korean Society for the Study of Obesity. J. Obes. Metab. Syndr. 32, 121–129 (2023).

60. Hassapidou, M. et al. European Association for the Study of Obesity Position Statement on Medical Nutrition Therapy for the Management of Overweight and Obesity in Adults Developed in Collaboration with the European Federation of the Associations of Dietitians. Obes. Facts 16, 11–28 (2023).

61. Agrawal, S. et al. Inherited basis of visceral, abdominal subcutaneous and gluteofemoral fat depots. Nat. Commun. 13, 3771 (2022).

62. Jurgens, S. J. et al. Rare coding variant analysis for human diseases across biobanks and ancestries. Nat. Genet. 56, 1811–1820 (2024).

63. Koenig, Z. et al. A harmonized public resource of deeply sequenced diverse human genomes. Genome Res. 34, 796–809 (2024).

64. Schizophrenia Working Group of the Psychiatric Genomics Consortium et al. LD Score regression distinguishes confounding from polygenicity in genome-wide association studies. Nat. Genet. 47, 291–295 (2015).

65. Privé, F., Arbel, J. & Vilhjálmsson, B. J. LDpred2: better, faster, stronger. Bioinformatics vol. 36 5424–5431 (2020).

66. Band, G. & Marchini, J. BGEN: a binary file format for imputed genotype and haplotype data. Preprint at 10.1101/308296 (2018).

67. Chang, C. C. et al. Second-generation PLINK: rising to the challenge of larger and richer datasets. GigaScience 4, 7 (2015).

